# Intervention Development Protocol for a novel, co-produced, sexually transmitted infection partner notification intervention for men who have sex with men

**DOI:** 10.1101/2020.10.21.20209049

**Authors:** Jean M McQueen, Melvina Woode Owusu, Fiona Mapp, Claudia S Estcourt, Merle Symonds, Alison R Howarth, Rak Nandwani, Susannah Brice, Alex Comer, Paul Flowers

**Author notes:** **CORRESPONDENCE** Paul Flowers.

## Abstract

**Introduction:** The number of bacterial sexually transmitted infections diagnosed among men who have sex with men (MSM) continues to rise annually. Innovative public health interventions are needed to address this. Partner notification (PN), is important in reducing STI transmission by identifying, testing and treating the sex partners of people with STIs. Outcomes of PN in MSM are sub-optimal; some MSM with STIs report high numbers of “one-off” sex partners (where sex occurs on one occasion only) who appear to contribute disproportionately to community transmission but are poorly reached by current PN interventions.

**Aims/Objectives:** This paper describes the protocol for development of a novel, co-produced, multi-level, PN intervention for MSM with “one-off” partners. The process described will ensure the intervention is evidence-based, theoretically informed and acceptable to users, service providers, commissioners and those with community interest.

**Methods and Analysis:** Our three-phase approach draws on the revised Medical Research Council (MRC) guidance for developing and evaluating complex interventions. First, we combine evidence synthesis with stakeholder engagement to understand the barriers and enablers to PN to co-produce preliminary intervention ideas. Next, we further develop our intervention ideas and adapt our emerging programme theory by collecting detailed data through focus groups and interviews with purposively sampled stakeholders. Data analysis using the theoretical domains framework and the behaviour change wheel will detail the relationship between putative causal mechanisms and optimal intervention components involved in enhancing PN amongst MSM. Finally, we refine our programme theory, map and clarify our intervention and its intersecting components. We will share our intervention with a panel of expert clinicians, third sector organisations and a lay audience of MSM to detail a co-produced PN intervention.

**Outcome:** Co-produced intervention and programme theory suitable for testing in a future feasibility study.

**Ethics and dissemination:** This protocol received ethical approval from Glasgow Caledonian University HLS/NCH/19/059. Findings will be published with open access licenses.

**SUMMARY:** Partner notification for men who have sex with men is suboptimal this paper describes a protocol to develop a co-produced multi-level partner notification intervention.

## INTRODUCTION

There is need for new, effective public health interventions to address increasing numbers of diagnosed sexually transmitted infections (STIs) particularly among men who have sex with men (MSM) given finite service capacity, rising demand and budgetary constraints [1]. In England, MSM account for over three quarters of syphilis diagnoses and just over half of reported gonorrhoea diagnoses [2] with a call for action made by the WHO setting a 90% reduction target in the global incidence of gonorrhoea by 2030 [3]. Curbing the incidence of STIs by reducing transmission is increasingly important given the sustained increase in several reported bacterial STIs among MSM internationally and evidence of emerging anti-microbial resistance in key pathogens such as *Neisseria gonorrhoea* and *Mycoplasma genitalium* [4].

Partner notification (also known as contact tracing) aims to reduce transmission by identifying, testing and treating the sex partners of people diagnosed with STIs [5]. PN for MSM remains poorly understood and optimal intervention approaches for disrupting the onward transmission of STIs among one-off sex partners are largely unknown [6]. MSM with bacterial STIs, such as gonorrhoea and syphilis, are likely to be at high risk of HIV infection. Improved PN creates an opportunity not just to improve health (by treating an undiagnosed STI) but also to prevent transmission, and to support contact with health professionals with opportunities to prevent HIV infection through PrEP. High numbers of STIs among MSM, and the extensive number of STIs which remain undetected, support the need and development of innovative PN methods able to reach anonymous, possibly high risk partners [7,8]. PN with MSM can be challenging owing to high numbers of anonymous sex partners, “one-off” partners (i.e. a one-off sexual encounter with little or no anticipation of sex again and little or no romantic connection) who may be particularly important in sustaining community transmission [7].

The places in which MSM meet sex partners have changed with a shift away from sex on premises venues to “digital dating”/digitally mediated, via geo-spatial applications (apps) and websites [7]. The use of smart phones and dating apps over the last decade has facilitated rapid partner change with increased opportunity for one-off encounters and STI transmission [9]. Increased digital dating may mean that previously uncontactable “one-off” sex contacts may be contactable. This brings opportunities for novel PN interventions for those who may contribute significantly to community STI transmission. PN targeting casual partners, rather than regular or live-in partners, prevents more secondary transmissions per partnership; although it is also more challenging and resource intensive, the public health benefit is greater [10].

PN for one-off partners can be particularly complex because, from the index patient’s perspective (the person diagnosed with the STI), there may be little motivation to engage in PN, there may be perceived fear, shame or guilt related to the STI and/or concern around negative reaction of their one-off partner/s [11]. Lack of emotional connection means MSM may not prioritise PN for those with whom they do not plan to see again and the practicalities associated with lack of contact details for one-off partners, adds a further layer of complexity. There is also additional service burden as PN for this partnership type can be labour intensive for providers, leading to resource pressures [12].

Furthermore, when working with health care professionals with limited resources to achieve PN outcomes, one-off partners may not be prioritised and PN with partners who are easily contactable and likely to have sex with the index again are more likely to become the primary focus of PN work. This creates complexity which demands a multi-level inclusive approach focused on what shapes PN with one-off partners and what could be done about it. This multi-levelled approach includes the role of the wider systems, key settings, actors and interactions that could be involved. For example, whilst smart phones and dating apps have contributed to changes in sexual practices (making it easier to meet others for casual sex) the use of digital technology may also have public health value. Digital technology allows communication of messages quickly and inexpensively offering digital and online approaches to inform partners who may traditionally have been more difficult to reach.

## INTERVENTION DEVELOPMENT

Our intervention development forms part of the Limiting Undetected Sexually Transmitted Infections to RedUce Morbidity (LUSTRUM) programme, funded by the National Institute for Health Research (NIHR) (Reference Number RP-PG-0614-20009). Our aim is to develop culturally appropriate cost-effective interventions for PN amongst MSM focusing on ‘one-off contacts’ where the potential for onward transmission of sexually transmitted infections (STIs) is likely to be greatest [10].

Intervention development processes are often poorly reported with wide variations in both the design, methods and language used [13]. Over the last 15 years’ intervention development has shifted from simplified processes based on the natural sciences or simply professional intuition, towards approaches that directly address issues of complexity and scientific enquiry [14]. Complexity may relate to intervention components, the health issues under study and or the behavioural systems the interventions seek to address, or indeed to the complexity of the contexts in which these interventions are implemented. With growing interest in intervention development, details of optimal approaches combined with theoretical frameworks are likely to be of increasing interest to those looking to plan and develop complex interventions. Often, information published about design is insufficient in detail and therefore doesn’t support replication. In addition, a paucity of evidence about the intervention development process and a lack of evaluation of resulting interventions means that it isn’t always possible to develop evidence-based interventions [15]. Recent calls to address these issues have identified the need to publish transparent intervention development protocols *prior* to intervention development [16].

Drawing on evidence informed principles such as the MRC guidance for developing and evaluating complex interventions, our project takes an iterative, creative, and forward looking progressive approach towards intervention development [16,17]. Meticulous development of all stages of complex interventions is needed to increase the likelihood of interventions being widely adopted and sustained [16]. The updated MRC guidance, under consultation in 2019, reinforces the need for exploration of the evidence, to develop theory led questions. This updated guidance considers the wider context in which interventions operate, the need to include meaningful stakeholder engagement and inclusion of programme theory and logic models to map key intervention processes and outcomes [18]. Creating a bridge between the different social worlds of academics, service users, clinicians and public health leads in intervention development research requires effective, clearly mapped strategies [13]. Increasingly, guidance highlights the importance and need for collaboration between researchers, front line practitioners and communities drawing on the expertise of key stakeholders through co-production [18,13]. During the co-production and intervention development process visual representation of interventions and how they are anticipated to work is one method to foster communication and collaboration required [19]. This can be achieved through the creation of co-produced iteratively developed programme theory [19]. The use of visual diagrams such as logic models helps structure focussed engagement, through an iterative yet robust process, which involves collaboratively evaluating intervention components with those who are likely to deliver and receive the interventions [20].

## BEHAVIOUR CHANGE THEORY FOR INTERVENTION DEVELOPMENT

Enhanced partner notification for MSM and their one-off partners can only be achieved by changing the behaviour of a series of actors implicated within the process (potentially MSM, health care professionals, those providing and managing MSM social networks and dating sites). Behavioural theory and accompanying theoretical frameworks provide a systematic process that can enhance intervention development [21,22] and provide a common language to describe intervention content. It should also consider the key settings in which the intervention will be implemented (i.e., the social world of practice) and finally, the health and social care systems in which co-produced - interventions can be implemented and embedded over time, alongside other health promoting activities.

Understanding behaviour and use of behaviour change theory is required to maximise the likelihood of intervention effectiveness [23,24]. The Theoretical Domains meta theoretical framework (TDF) will be used as a way of describing the causal mechanisms that our intervention moderates. In recent years, international research has emerged using the theoretical domains framework (TDF) to assess behavioural problems and develop behaviour change interventions targeting clinical staff [18,25]. The TDF combines 128 constructs from 33 behaviour change theories into 14 theoretical domains used for intervention design and to help understand the behaviour change process [22]. The behaviour change wheel (BCW) [30] will be used to provide a common language to describe our intervention content, ranging from intervention functions to individual behaviour change techniques (BCTs).

**AIM-** to develop a novel, co-produced, STI partner notification intervention for men who have sex with men which is evidence-based, theoretically informed and acceptable to users, healthcare professionals, digital technology and dating app providers.

## METHODS

### OVERARCHING DESIGN

We will adopt a multi-phase pluralistic approach to intervention development (figure 1). Our approach is based upon a sequential design that both builds and focuses on useful learning. Figure one shows the three phases of intervention development. Broadly speaking, **Phase 1:** combines evidence synthesis with initial stakeholder expertise, to develop preliminary programme theory describing how a co-ordinated, multi-level intervention compromised of various components may be able to reduce the onwards transmission of STIs amongst MSM (table 1 phase one objectives). **Phase 2:** takes these preliminary ideas and systematically explores them further through behaviourally informed qualitative research. It ends with a refined programme theory. In **Phase 3:** we will return to our stakeholders and refine the programme theory once more, ensuring that the proposed intervention and its interdependent components are acceptable, implementable and fit for purpose.

**Table 1:** Appease Criteria (Mitchie et al 2011)

|  |  |
| --- | --- |
| Acceptability | How far is it acceptable to all key stakeholder? |
| Practicability | Can it be implemented as designed within the intended context, material and human resources? |
| Effectiveness | How effective and cost effective is it in achieving desired objectives in the target population? |
| Affordability | How can it be afforded when delivered at the scale intended? |
| Side-effects | How far does It lead to adverse or beneficial outcomes? |
| Equity | How far does it increase or decrease differences between advantaged and disadvantaged sectors of society? |

**Figure 1:**
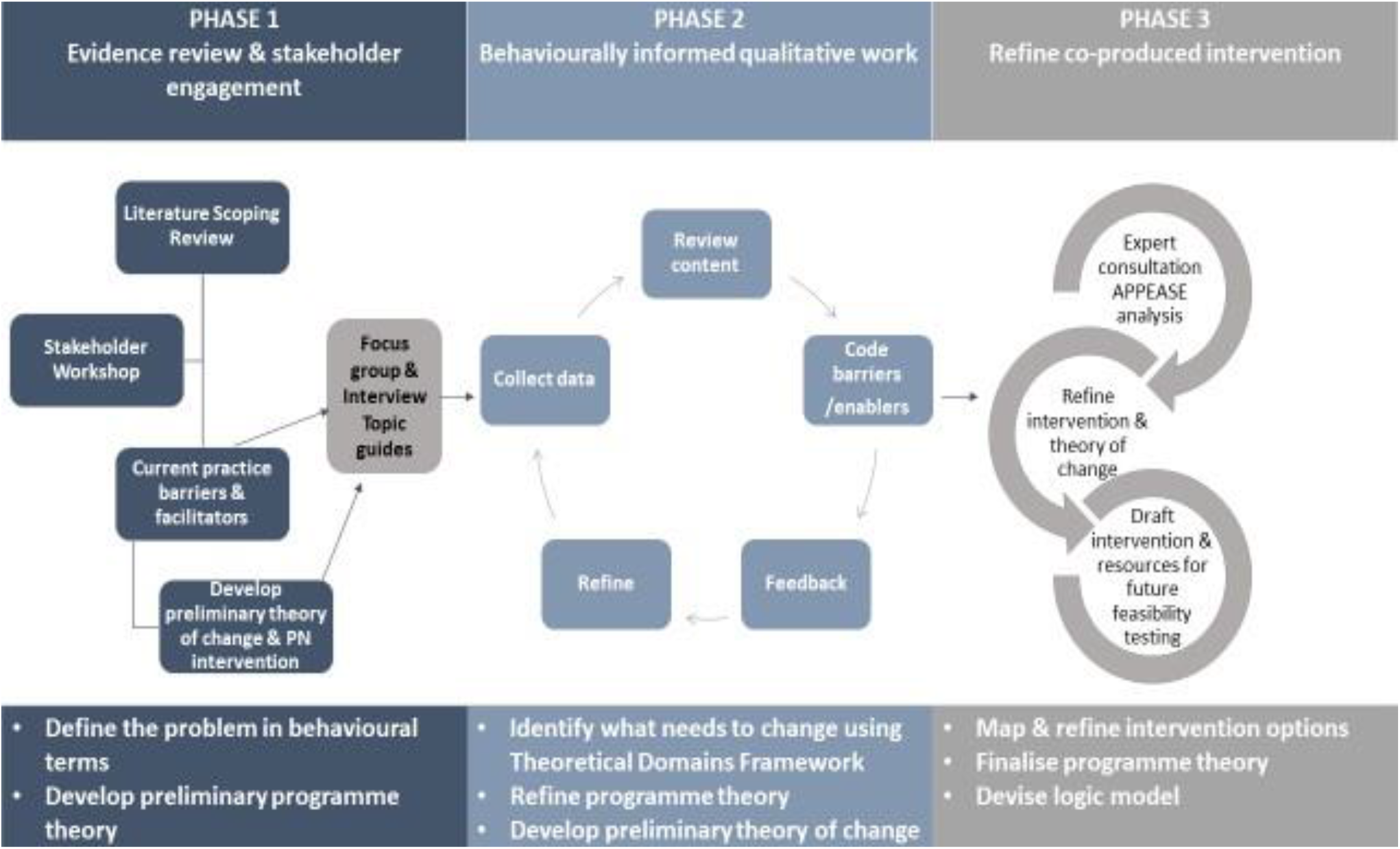
Developing STI partner notification interventions for men who have sex with men and their one-off partners.

#### Phase 1: Evidence synthesis and the collation of stakeholder-led preliminary intervention ideas

To ensure rigour in our scoping review, we will use the approach developed by Arksey and O’Malley [26] and refined by Levac et al [27] and the Joanna Briggs Institute [28]. The searches will be limited to studies implementing PN interventions aimed at MSM. All peer-reviewed studies written in English published from the year 2000 to date will be included. The following databases will be searched CINAHL, ProQuest, PsycInfo, EMBASE, AMED, ASSIA, PubMed. We will undertake lateral searches from the reference lists of included studies. Two reviewers will independently screen titles and abstracts for inclusion, followed by screening full text of potentially relevant articles to determine final inclusion. Any discrepancies will be discussed. Types of PN interventions will be mapped to inform stakeholder discussion topics and our future intervention development.

Next, we will host a stakeholder workshop exploring in detail current PN practice, barriers and facilitators to PN, intervention ideas (derived from earlier scoping review) and opportunities to change the current state of affairs and wider systems in which they operate. The workshop will involve a diverse range of stakeholders including MSM, health care providers, service commissioners, public health leads, third sector organisations, dating app providers, technology providers and researchers. We anticipate this will include around 50 participants who will collaboratively explore PN for one-off partners in small groups, comprising a mix of different types of stakeholders in each group. Discussion will focus on generating ideas about who needs to do what, where and when to improve PN in MSM and one-off partners. Group discussions will be coordinated by facilitators who will encourage creative and ‘blue sky thinking’, exploring novel intervention ideas which are mapped to 1) what MSM can do 2) what HCPs can do, 3) what dating app providers can do and 4) what a combination of these groups working together can do. Information gathered from our scoping review and stakeholder engagement event will be used to further develop our programme theory for intervention development detailing how the intervention is proposed to work, showing the mechanisms by which our intervention influences the proposed outcomes [29].

##### Anticipated outputs

□ **Co-produced broad intervention ideas to take forward into phase 2**
□ **Stakeholder-led insights into multi-levelled barriers and facilitators to PN for MSM with one-off partners with problems defined in behavioural terms**
□ **Initial programme theory (to be refined in latter phases)**

Following phase 1, uncertainties are likely to remain as to the acceptability of our preliminary intervention ideas to those likely to use them, those not involved in our stakeholder event, and the barriers and enablers to these intervention ideas being implemented. These uncertainties will be explored in phase 2 through focus groups and interviews.

#### Phase 2: Behaviourally informed qualitative work, stakeholder focus groups and interviews

Phase 2 objectives are to:

1. Understand in detail the behaviours involved in operationalising the ideas developed within phase 1 using stakeholder focus groups and interviews focused on the barriers and facilitators to their uptake and implementation.
2. Use the behaviour change wheel (BCW figure 2) to detail the relationship between causal mechanisms implicated in uptake, implementation and optimal intervention components.
3. Refine the intervention and further develop the programme theory in light of what has been learned consulting with the range of stakeholders who could be involved in PN interventions (e.g., health care professionals, non-government organisations, MSM, digital technology providers) gathering information to tailor the PN intervention to the target population and context. Thus, maximising acceptability and reducing potential implementation problems later.

**Figure 2:**
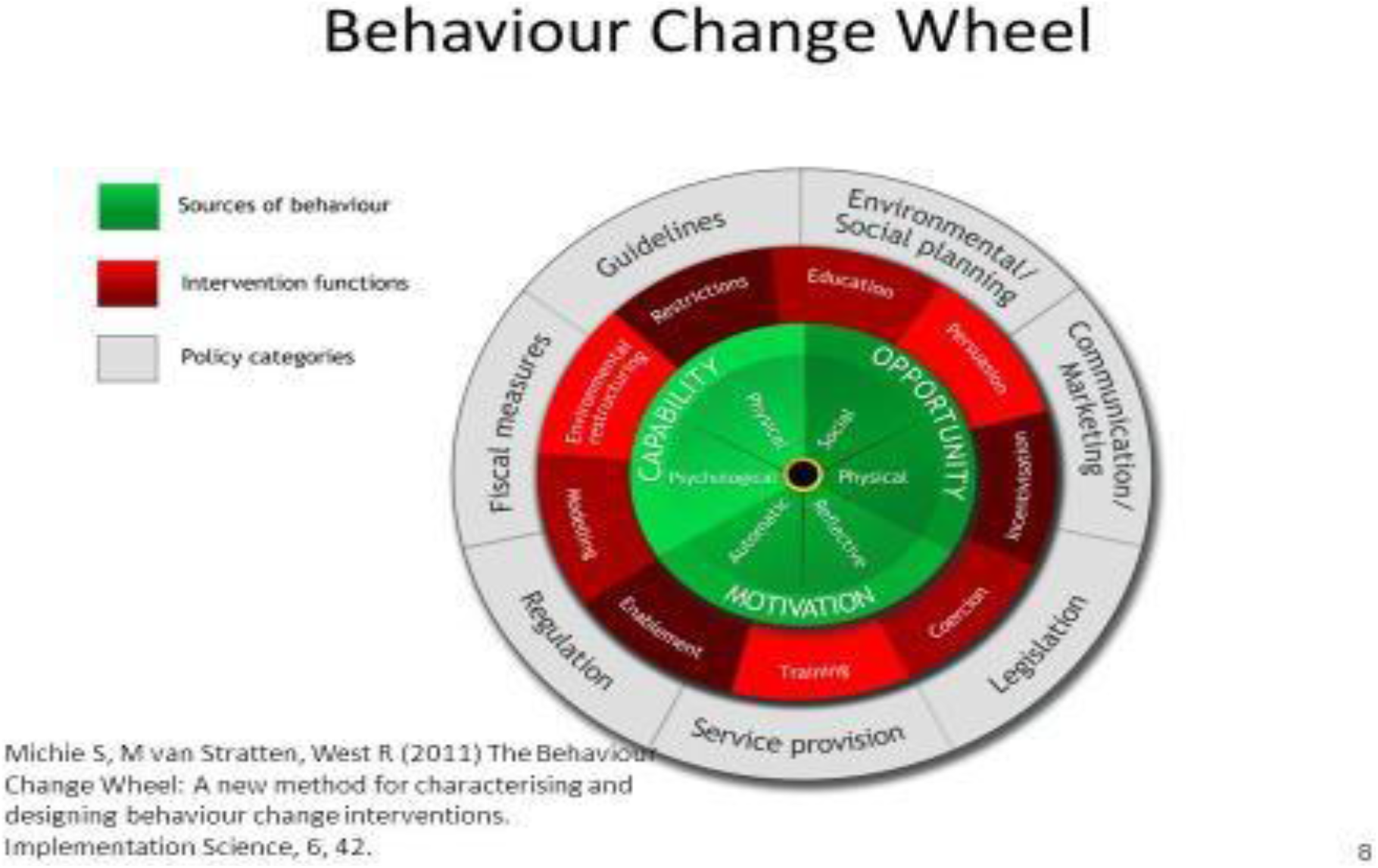
Behaviour change wheel.

During a series of focus groups and interviews we will present and discuss the broad intervention ideas from the stakeholder event in detail with a clear focus on the barriers and facilitators to uptake and implementation. This will involve around 40 purposely sampled stakeholders i.e. MSM, digital technology providers, expert clinicians, non-government organisations and any other additional stakeholders identified throughout the process. Separate focus groups will be conducted for each stakeholder group i.e. MSM, HCPs, NGOs with dating app providers offered individual interviews.

The topic guides for both the interviews and focus groups will be informed by our visual and high level summary mapped in our programme theory created in phase 1 and include intervention ideas informed by the scoping review and stakeholder workshop. These broad intervention ideas will form the basis of focus groups and telephone interviews with MSM, dating app providers and expert clinicians. Purposive sampling will be used to identify up to 28 MSM, 3 dating app providers, 6 expert clinicians and 3 non-government organisations offering MSM sexual health advice. All interviews and focus group data will be digitally recorded, transcribed and thematically analysed with a mix of inductive learning from our participants combined with deductively adding more depth and data around the barriers and facilitators to uptake and implementation of our intervention ideas.

Different yet complementary analytic approaches will be used. Our analysis of the problems associated with PN from phase 1 (defined in behavioural terms) will be used to identify the behaviours that need to change to improve PN. The TDF domains will be used to identify and explore the barriers and facilitators to change. The BCW will be used to identify the interacting behaviours from key actors requiring change. The intervention functions that form the third layer of the BCW will support practical intervention recommendations, intervention content and implementation options to enhance PN for MSM with one-off partners. The construct domains, themes and the barriers and enablers to the novel PN intervention will be mapped to the BCW (figure 2). Analyses of the interview transcripts will be used to identify barriers and enablers to the proposed PN intervention, ways in which PN could be enhanced, who needs to do what with whom and what might work for whom. Phase 2 ends with a refined programme theory with detailed and specific intervention ideas which we will take forward into phase 3.

##### Anticipated outputs

□ **Barriers and facilitators to change identified and coded using TDF**
□ **Behaviours requiring change specified with potentially useful intervention functions**
□ **Practical intervention recommendations specified to include named behaviour change techniques**

Following phase 2, uncertainties are likely to remain as to how the specific intervention components will work together to enhance PN, the predicted impact and economic costs of the intervention, and whether it is feasible, practicable and able to be implemented in the real world.

#### Phase 3: Refined programme theory with detailed PN intervention ready for feasibility testing

The intervention development process concludes with Phase 3, where we will refine our programme theory, and detail how specific intervention components are expected to enhance PN. Here we map out the detailed PN intervention ready for feasibility testing in a subsequent trial. This phase will be designed to facilitate ongoing stakeholder engagement with the intervention development process, and establish networks through which a preliminary feasibility and ‘real world’ implement-ability trial could be conducted.

We will share details of the intervention ideas, iteratively developed programme theory further exploring if these are acceptable, feasible and implementable in the ‘real world’. Here we will present our intervention ideas to health care professionals, clinical experts and academic partners. The APPEASE criteria (table 1) will be used to determine if the intervention is affordable, practical, effective, acceptable, has **side effects** and drives inequalities in experiences or outcomes or prevents the intervention being distributed/equally [30].

Use of the APPEASE criteria will sharpen the PN intervention moving from a long list of intervention ideas to more specific and focused intervention, ready for feasibility testing. Phase 3 ends with a refined programme theory and prototype evidence based, co-produced PN intervention. Following this it is likely that a period of testing the intervention content, materials and delivery methods will be necessary to identify early issues with acceptability, feasibility and other potential teething problems to allow these to be addressed prior to formal piloting and evaluation in a feasibility study.

##### Anticipated outputs

□ **a refined programme theory**
□ **co-produced PN intervention tailored to MSM**

The study will take 12 months (table 2). At the end of this work, the feasibility of the new PN intervention will be tested in a pilot study as we move into the feasibility stage of MRC guidance on development of complex interventions [18].

**Table 2:**
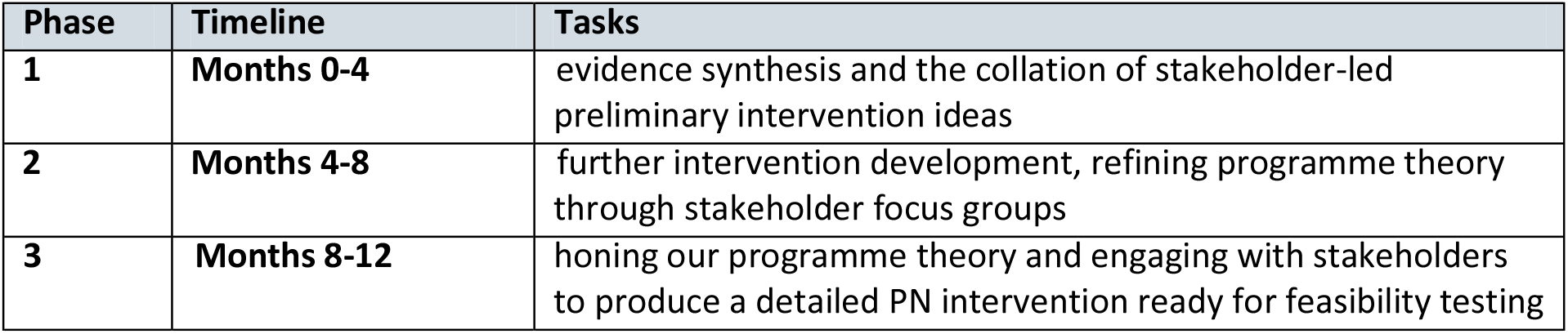
Timeline.

| Phase | Timeline | Tasks |
| --- | --- | --- |
| **1** | **Months 0-4** | evidence synthesis and the collation of stakeholder-led preliminary intervention ideas |
| **2** | **Months 4-8** | further intervention development, refining programme theory through stakeholder focus groups |
| **3** | **Months 8-12** | honing our programme theory and engaging with stakeholders to produce a detailed PN intervention ready for feasibility testing |

### COMPARISON WITH EXISTING LITERATURE

The authors believe that this programme of work with its focus on intervention development around PN for MSM is novel in the UK and will therefore make an original research contribution. Reflections on the process should provide a useful insight for future research around intervention development. Due to COVID-19 pandemic, some of the Phase 2 and 3 stakeholder engagement will be virtual and this represents a novel engagement approach.

### IMPLICATIONS FOR PRACTICE

This project has the potential to impact and benefit MSM and their sexual networks, health care professionals who deliver PN and wider society by helping prevent STIs and mitigate associated ill health. The findings have the potential to enhance PN changing attitudes and behaviours of MSM, providing health care professionals with much needed guidance, a framework for practice to improve PN and support evidence based, tailored PN. Broader public health impact would also be expected through reductions in the onward transmission of STIs. The intervention development process should provide useful practical insights and methodological contributions for future research around intervention development.

## Data Availability

This is a protocol data collection has not yet concluded and data that support the findings of this study will be available from the corresponding author, [Paul Flowers], upon reasonable request.

https://www.lustrum.org.uk/

## ACKNOWLEDGMENTS

The wider LUSTRUM research team: Christian Althaus, Jackie A Cassell, Andrew Copas, Nicola Low, Catherine H Mercer, Chidubem (Duby) Ogwulu, John Saunders, Tracy Roberts, Sonali Wayal

## AUTHOR CONTRIBUTIONS

CSE conceived the intervention development stream of the LUSTRUM programme, JMQ and PF conceptualised this programme, FM, MWO, MS, AH, JMQ, PF supported and contributed to data collection through stakeholder engagement, JMQ and PF analysed data, CE, RN, SB, AC contributed to intervention development and importance of study. JMQ and PF drafted this manuscript with input from all authors to finalise before submission.

## FUNDING

LUSTRUM is a five-year programme of research funded by the National Institute for Health Research (NIHR) under its Programme Grants for Applied Research (Reference Number RP-PG-0614-20009).

## ETHICAL APPROVAL

The relevant ethical approvals have been sought for phase 2 interviews and focus groups with key stakeholders. Ethical approval for focus groups and interviews with MSM and digital technology providers has been granted from Glasgow Caledonian University Ethics Committee (HLS/NCH/19/013).

## Notes

**FUNDING** This work was supported by the National Institute for Health Research (NIHR) Reference Number RP-PG-0614-20009

**CONFLICTS OF INTEREST** The authors declare no conflicts of interest.

### Competing Interest Statement

The authors have declared no competing interest.

### Clinical Trial

Protocol

### Author Declarations

This protocol received ethical approval from Glasgow Caledonian University HLS/NCH/19/059.

